# A Paradoxical Screening Serological Assay for the Diagnosis of Whipple’s Disease (infection with *Tropheryma whipplei*)

**DOI:** 10.1101/2021.01.15.21249681

**Authors:** K.C. Liew, Chelsea Nguyen, Nilakshi T Waidyatillake, John Stenos, Aaron Walton, Eugene Athan, Owen Harris, Stephen Graves

**Affiliations:** Department of Microbiology, Australian Clinical Labs, Geelong Laboratory, Geelong 3220, VIC, Australia; Australian Rickettsial Reference Laboratory, University Hospital Geelong, Geelong 3220, VIC, Australia; Deakin University, School of Medicine, Geelong 3220, VIC, Australia; Barwon Health, University Hospital Geelong 3220, VIC, Australia; Allergy and Lung Health Unit, Centre for Epidemiology and Bio statistics, Melbourne School of Population and Global Health, The University of Melbourne, VIC, Australia

**Author notes:** Address correspondence to K. C. Liew,. Alternative Corresponding author: Prof Stephen Graves. Author order was determined based on seniority and contribution to this work. This is contribution from the Australian Rickettsial Reference Laboratory in evaluating the screening serological assay for the diagnosis of Whipple’s Disease.

## Abstract

Whipple’s disease (WD) is a rare infection due to *Tropheryma whipplei*. Following *in-vitro* cultivation of *T. whipplei*, an indirect-immunofluorescence serological assay (IFA) was developed. We tested the hypothesis that this assay could be used to either identify WD patients, or rule out WD, in patients in whom the diagnosis is being considered, based on the antibody titers of their IgM and IgG antibody responses. In this small study fourteen WD patients and 22 healthy volunteers’ sera were obtained from across Australia. All specimens were coded and de-identified before testing. A patient with an IgG antibody titer of ≤1:16 may have WD [sensitivity 57% (8/14) and specificity close to 100% (22/22)]. High IgM antibody titers (≥1:256) were more common in WD patients [sensitivity 50% (7/14) and specificity 86% (19/22)] than in controls. The area under Receiver-Operator-Characteristic curve for IgG in the IFA assay was 0.84 (95% CI 0.69-1.00). At an IgG antibody titer of ≤1:16 the Youden’s index was 0.57. WD patients’ under-produce IgG antibody to *T*.*whipplei* but are more likely to over-produce IgM antibodies. This screening IFA serological assay may be clinically useful in detecting those with a possible diagnosis of WD. Patients with an IgG antibody titer of ≤1:16 and an IgM antibody titer of ≥1:256 may have WD and should proceed to a tissue biopsy and PCR for confirmation. Further validation of this assay, by increasing the sample size, by testing it in patients with non-WD disease and trialing in other countries should be undertaken.

## INTRODUCTION

Whipple’s disease (WD) is a rare infection with *Tropheryma whipplei* with an estimated prevalence of 1.1 to 9.8 cases per million inhabitants and an annual incidence of 1 to 6 per 10 million inhabitants (1-3). In small case series from Germany, Spain and north-western Italy, the maximum incidence of WD was in persons between 40-60 years of age, of whom 72-96% were middle-aged Caucasian men (2-7). In contrast to these findings, a large retrospective population-based study carried out between 2012 and 2017 in the United States (US) found that men and women were affected at similar rates and that WD was more common in people aged greater than 65 years (3).

The clinical spectrum of WD is wide (1,5,8,9). In classical WD, infection with *T. whipplei* probably occurs years before the first clinical manifestations, which are most often chronic diarrhea or migratory arthralgia (1,9). In localized disease, *T. whipplei* can infect the joints, bones, brain, heart, skin, eyes, lymph nodes, lung and/or kidney (1,8,9). Other associated syndromes include acute, self-limited gastroenteritis and pneumonia (1,9). A broad spectrum of clinical severity has been observed, ranging from asymptomatic carriage (5,8) to organ failure and death. Fatal outcome is associated with delayed diagnosis as well as misdiagnosis as an autoimmune/inflammatory disorder with consequent immunosuppressive treatment (9).

It has been observed that patients with confirmed WD frequently have reduced or undetectable *T. whipplei*-specific IgG antibody levels. We have hypothesized that WD patients may not be able to mount an adequate IgG antibody response. Persons who do not go on to develop clinical WD, by contrast, appear to mount a strong IgG antibody response when exposed to *T. whipplei*. While healthy individuals not yet exposed to the bacterium (mostly children and young adults) are necessarily IgG seronegative, they respond normally when exposed. The aim of this study was to compare the sera of patients with confirmed WD to healthy individuals without WD. We set out to evaluate the diagnostic utility of a *T. whipplei* screening IFA serology assay as a tool to distinguish patients with WD from those without.

## PATIENTS, MATERIALS AND METHODS

### Study Design

A small case comparative study of *T. whipplei* IFA was conducted from November 2019 until May 2020 at the Australian Rickettsial Reference Laboratory (ARRL), Geelong, Victoria, Australia, according to the *National Statement on Ethical Conduct in Human Research (2007)* guideline (Barwon Health ethics reference number: 19/135). Written informed consent was obtained from WD patients and healthy controls. All sera tested in this study were de-identified.

Participant selection criteria included English language proficiency and age > 18 years. Pregnancy, active malignancy, primary immunodeficiency and HIV with CD4 lymphocyte count < 200 were grounds for exclusion. Subjects were considered WD cases if they displayed consistent symptoms with supportive diagnostic test results, such as positive periodic acid-Schiff (PAS) staining, *T. whipplei*-specific immunohistochemistry staining or *T. whipplei* PCR of a biopsy specimen (10-12).

### Sample size

We employed a convenience sampling strategy, given the limited number of WD patients in Australia. In total, we recruited 36 participants. Fourteen patients with confirmed WD were recruited from across Australia. Twenty-two healthy controls were recruited locally.

### Diagnostic techniques

#### Culture and IFA slide preparation

A French strain of *T. whipplei* was cultivated in liquid axenic medium at 35°C for several weeks (13). Resultant *T. whipplei* cells were pelleted and resuspended in phosphate buffered saline (PBS). To optimize antigen concentrations for the final immunofluorescence assay (IFA), slides were prepared using doubling dilutions of *T. whipplei* antigen and subjected to sandwich IFA after incubation with a strongly reacting serum sample. Antigen aliquots were stored at −70°C until required. When thawed, they were spotted onto clean 3 x 10 well glass slides. After air drying, antigens were fixed with acetone for 5 minutes. Slides could then be stored at −70°C for later use.

### Indirect-immunofluorescence assay (IFA)

All sera from the WD patients and controls were tested by our in-house *T. whipplei* IFA. Known reactive and non-reactive sera were tested on the same slide as the assay samples. Initial 1:8 dilutions of test and control sera were prepared using 2% casein PBS buffer. Serial doubling dilutions from 1:8 to 1:4096 were prepared and tested by IFA. Dilute sera were spotted in duplicate onto pre-prepared *T. whipplei* slides. Slides were incubated for 30 minutes under humid conditions at 35°C and thereafter washed for 5 minutes in a 1:10 dilution of PBS and allowed to dry. Fluorescein-conjugated goat-anti-human antibodies, [1:100 anti-human IgA (KPL, Cat # 02-10-01), 1:100 anti-human IgM (KPL, Cat # 02-10-03) and 1:100 anti-human IgG (KPL, Cat # 02-10-02)] were applied, and slides were incubated, washed and dried as above. Fluorescence mounting fluid (Dako North America, Inc., USA) and a coverslip were applied. Microscopic evaluations of the IFA were carried out under 400x magnification using an LED Leica DMLS fluorescence microscope. The assay was repeated in duplicate for all sera. Antibody titers for IgM, IgG and IgA were defined as the highest serum dilution giving strong immunofluorescence.

### Sample collection

Of the 14 WD cases, eleven had sera already in storage. The remaining three cases had serum sample collection organized through their local doctors and pathology services. Healthy control sera were collected by the study doctor (KCL). Data such as gender, age range, IgM, IgG and IgA antibody titers to *T. whipplei* were recorded.

### Statistical analysis

Statistical comparisons of groups, by 2-sided Fisher exact test was done on the quantitative data. Sensitivity was defined as the proportion of WD patients correctly identified at a particular antibody cutoff (14). The definition for specificity was the proportion of healthy controls correctly identified as not having WD (15). The clinical accuracies of the *T. whipplei* IFA assays were examined by using Receiver-Operator-Characteristic (ROC) plots with GraphPad Prism version 8.0.2 (GraphPad Software, San Diego, CA, USA) (15). ROC area under the curve (AUC) were calculated as the fraction “correctly identified to be positive” and the fraction “falsely identified to be positive” determined according to researcher cutoff values for positive results (15). The Youden index, a function of sensitivity and specificity, was a commonly used measure of overall diagnostic effectiveness (16). This index ranged between 0 and 1, with values close to 1 indicating that the biomarker’s effectiveness was relatively large, and values close to 0 indicating limited effectiveness (16). Likelihood determined the probability that a test result was correct to the probability that the test result was incorrect (17).

## RESULTS

### Case presentations and demographics

The clinical presentations of the fourteen WD cases included in this study were varied. Half of them had infective endocarditis; three subjects displayed central nervous system manifestations; two had classic WD with intestinal malabsorption, and two had joint manifestations. All but one case subject was male. Ages ranged from 36-85 years old, with a mean age of 59 (Table 1).

**TABLE 1:**
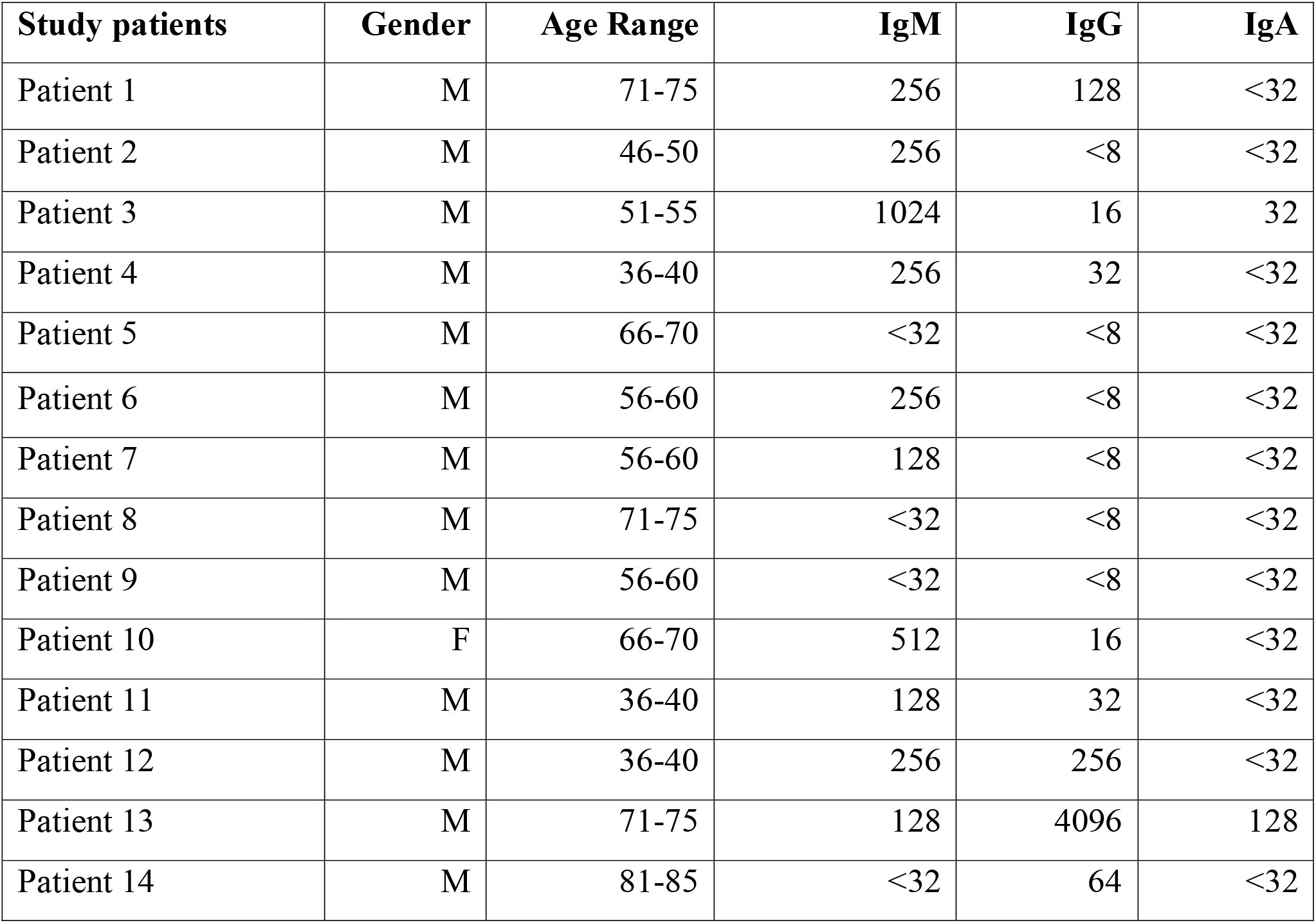
Whipple’s Disease patients’ indirect immunofluorescence assay antibody titers.

| **Study patients** | **Gender** | **Age Range** | **IgM** | **IgG** | **IgA** |
| --- | --- | --- | --- | --- | --- |
| Patient 1 | M | 71-75 | 256 | 128 | <32 |
| Patient 2 | M | 46-50 | 256 | <8 | <32 |
| Patient 3 | M | 51-55 | 1024 | 16 | 32 |
| Patient 4 | M | 36-40 | 256 | 32 | <32 |
| Patient 5 | M | 66-70 | <32 | <8 | <32 |
| Patient 6 | M | 56-60 | 256 | <8 | <32 |
| Patient 7 | M | 56-60 | 128 | <8 | <32 |
| Patient 8 | M | 71-75 | <32 | <8 | <32 |
| Patient 9 | M | 56-60 | <32 | <8 | <32 |
| Patient 10 | F | 66-70 | 512 | 16 | <32 |
| Patient 11 | M | 36-40 | 128 | 32 | <32 |
| Patient 12 | M | 36-40 | 256 | 256 | <32 |
| Patient 13 | M | 71-75 | 128 | 4096 | 128 |
| Patient 14 | M | 81-85 | <32 | 64 | <32 |

### Healthy controls’ demographics

Of the healthy controls, 6 of 22 (27%) were males with ages ranging from 20-75 years old (Table 2). Mean age of healthy controls was forty.

**TABLE 2:** Indirect immunofluorescence assay antibody titers for healthy controls

| **Healthy Controls** | **Gender** | **Age Range** | **IgM** | **IgG** | **IgA** |
| --- | --- | --- | --- | --- | --- |
| Control 1 | F | 56-60 | 64 | 256 | <32 |
| Control 2 | F | 41-45 | 128 | 256 | <32 |
| Control 3 | F | 41-45 | 64 | 256 | <32 |
| Control 4 | F | 36-40 | 128 | 128 | 32 |
| Control 5 | F | 41-45 | 32 | 32 | <32 |
| Control 6 | F | 46-50 | 64 | 512 | 128 |
| Control 7 | F | 46-50 | 256 | 256 | <32 |
| Control 8 | M | 51-55 | 64 | 256 | <32 |
| Control 9 | F | 26-30 | 512 | 64 | 256 |
| Control 10 | F | 41-45 | 32 | 512 | 32 |
| Control 11 | F | 31-35 | <32 | 2048 | <32 |
| Control 12 | F | 31-35 | 512 | 256 | <32 |
| Control 13 | F | 36-40 | 128 | 512 | <32 |
| Control 14 | M | 36-40 | <32 | 256 | <32 |
| Control 15 | M | 71-75 | <32 | 512 | <32 |
| Control 16 | F | 21-25 | 128 | 32 | 32 |
| Control 17 | M | 21-25 | 128 | 32 | <32 |
| Control 18 | M | 26-30 | <32 | 256 | <32 |
| Control 19 | M | 36-40 | <32 | 128 | 32 |
| Control 20 | F | 41-45 | 128 | 32 | <32 |
| Control 21 | F | 21-25 | <32 | 256 | <32 |
| Control 22 | F | 50-55 | <32 | 512 | <32 |

### IgG IFA

WD patients (n=14) (Table 1) were less able to produce IgG antibodies to *T. whipplei* compared with healthy controls (n=22) (Table 2) and shown by comparison in Table 3. This difference was significant when comparing antibody titers at 1:16, 1: 32, 1:64; 1:128 and 1:256 dilutions (*p <*.*05*).

**TABLE 3:** Serological comparison of IgG antibody titers in Whipple’s Disease (WD) patients and healthy controls; role in excluding WD in patients.

| **Antibody**<br>**Titers** | **Whipple's**<br>**Disease** | **Healthy**<br>**Controls** | **Sensitivity &**<br>**Specificity** | **p-value<sup>@</sup>** |
| --- | --- | --- | --- | --- |
| **IgG <256**<br>**(Test +)** | 12 | 7 | Sn: 86%<br>(95% CI 57-98%)<br>Sp: 68%<br>(95% CI 45-86%) | 0.0022 |
| **IgG ≥ 256**<br>**(Test -)** | 2 | 15 |  |  |
| **IgG < 128**<br>**(Test +)** | 11 | 5 | Sn: 79%<br>(95% CI 49-95%)<br>Sp: 77%<br>(95% CI 55-92%) | 0.0017 |
| **IgG ≥ 128**<br>**(Test -)** | 3 | 17 |  |  |
| **IgG < 64**<br>**(Test +)** | 10 | 4 | Sn: 71%<br>(95% CI 42-92%)<br>Sp: 82%<br>(95% CI 60-95%) | 0.004 |
| **IgG ≥ 64**<br>**(Test -)** | 4 | 18 |  |  |
| **IgG < 32**<br>**(Test +)** | 8 | 0 | Sn: 57%<br>(95% CI 29-82%)<br>Sp: 100%<br>(95% CI 85-100%) | 0.0001 |
| **IgG ≥ 32**<br>**(Test -)** | 6 | 22 |  |  |
| **IgG < 16**<br>**(Test +)** | 6 | 0 | Sn: 43%<br>(95% CI 18-71%) | 0.0015 |
| **IgG ≥ 16**<br>**(Test -)** | 8 | 22 | Sp: 100%<br>(95% CI 85-100%) |  |

<sup>@</sup> p-values are determined by Fisher's exact probability test; Sn: Sensitivity; Sp: Specificity

At a serum dilution of 1:256, 15 of 22 (68%) healthy controls had IgG to *T. whipplei* compared to only 2 of 14 (14%) WD cases (p<0.05). With an IgG titer cutoff set at less than 1:256, sensitivity and specificity of the IFA were 86% (95% CI 57-98%) and 68% (95% CI 45-86%) respectively (Table 3).

All twenty-two healthy controls had detectable *T. whipplei* IgG at a serum dilution of 1:32, compared with 6 of 14 (43%) WD cases (p<0.05). While the sensitivity was only 57% at this cutoff, specificity approached 100% (95% CI 85-100%). The area under the Receiver-Operator-Characteristic curve for IgG was 0.84 (95% CI 0.69 – 1.00). At an IgG antibody titer cutoff < 1:32, the Youden’s index was 0.57 (Figure 1).

**FIG 1:**
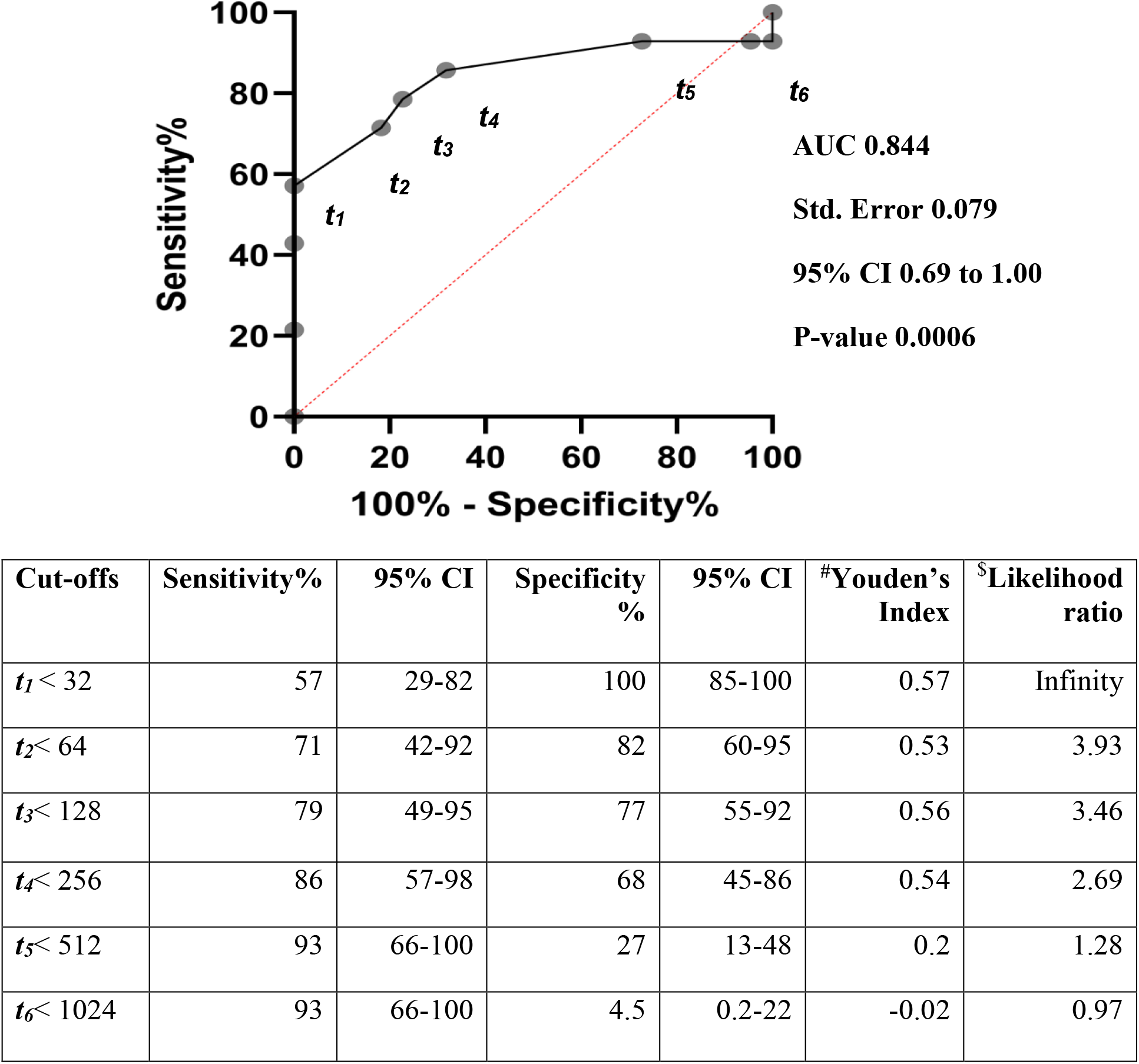
Comparison of IgG antibody titers in Whipple’s Disease patients and healthy controls: ROC. ^#^Youden’s index represents a summary measurement of the ROC for the accuracy of a diagnostic test with ordinal or continuous endpoints. ^$^Likelihood ratio is basically a ratio of the probability that a test result is correct to the probability that the test result is incorrect.

### IgM IFA

IgM antibodies to *T. whipplei* were detected in a higher proportion of WD cases than in healthy controls (*p = 0*.*03*) (Table 4). With the titer cutoff set at 1:256, sensitivity and specificity for the IgM IFA were 50% (95% CI 23-77%) and 86% (95% CI 65-97%) respectively. The area under the ROC curve was 0.65 (95% CI 0.45-0.86). Using this cutoff, Youden’s index was 0.36 (Figure 2).

**TABLE 4:** Serological comparison of Whipple’s Disease patients and healthy controls at an IgM antibody titer of ≥ 256

| **Antibody**<br>**Titers** | **Whipple's**<br>**Disease** | **Healthy**<br>**Controls** | **Sensitivity &**<br>**Specificity** | **p-value<sup>@</sup>** |
| --- | --- | --- | --- | --- |
| **IgM $\geq 256$**<br>**(Test +)** | 7 | 3 | Sn: 50%<br>(95% CI 23-77%) | 0.03 |
| **IgM $< 256$**<br>**(Test -)** | 7 | 19 | Sp: 86%<br>(95% CI 65-97%) |  |

<sup>@</sup> p-values are determined by Fisher's exact probability test; Sn: Sensitivity; Sp: Specificity

**FIG 2:**
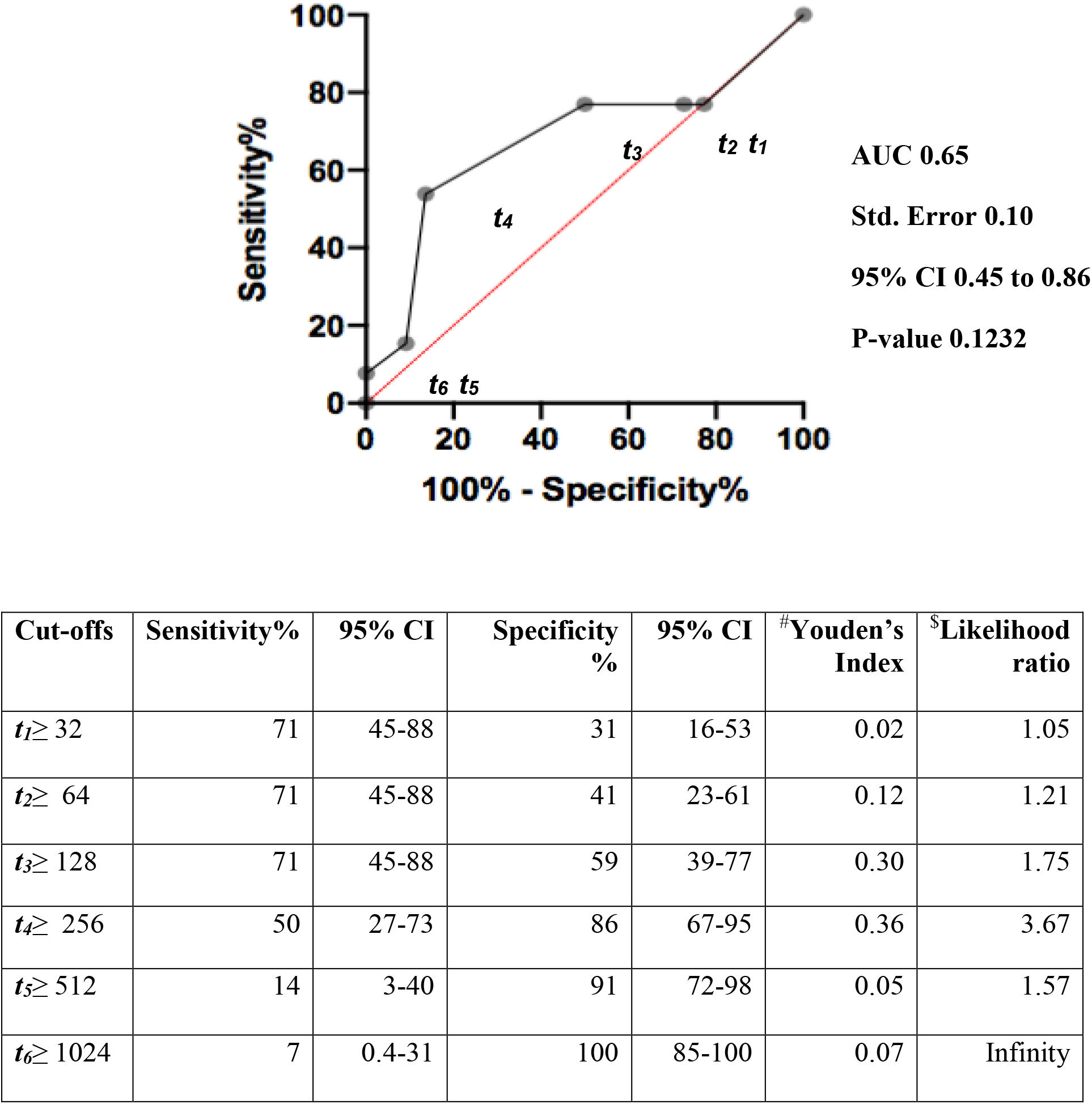
Comparison of IgM antibody titers in Whipple’s Disease patients and healthy control: ROC. ^#^Youden’s index represents a summary measurement of the ROC for the accuracy of a diagnostic test with ordinal or continuous endpoints. ^$^Likelihood ratio is basically a ratio of the probability that a test result is correct to the probability that the test result is incorrect.

**FIG 3:**
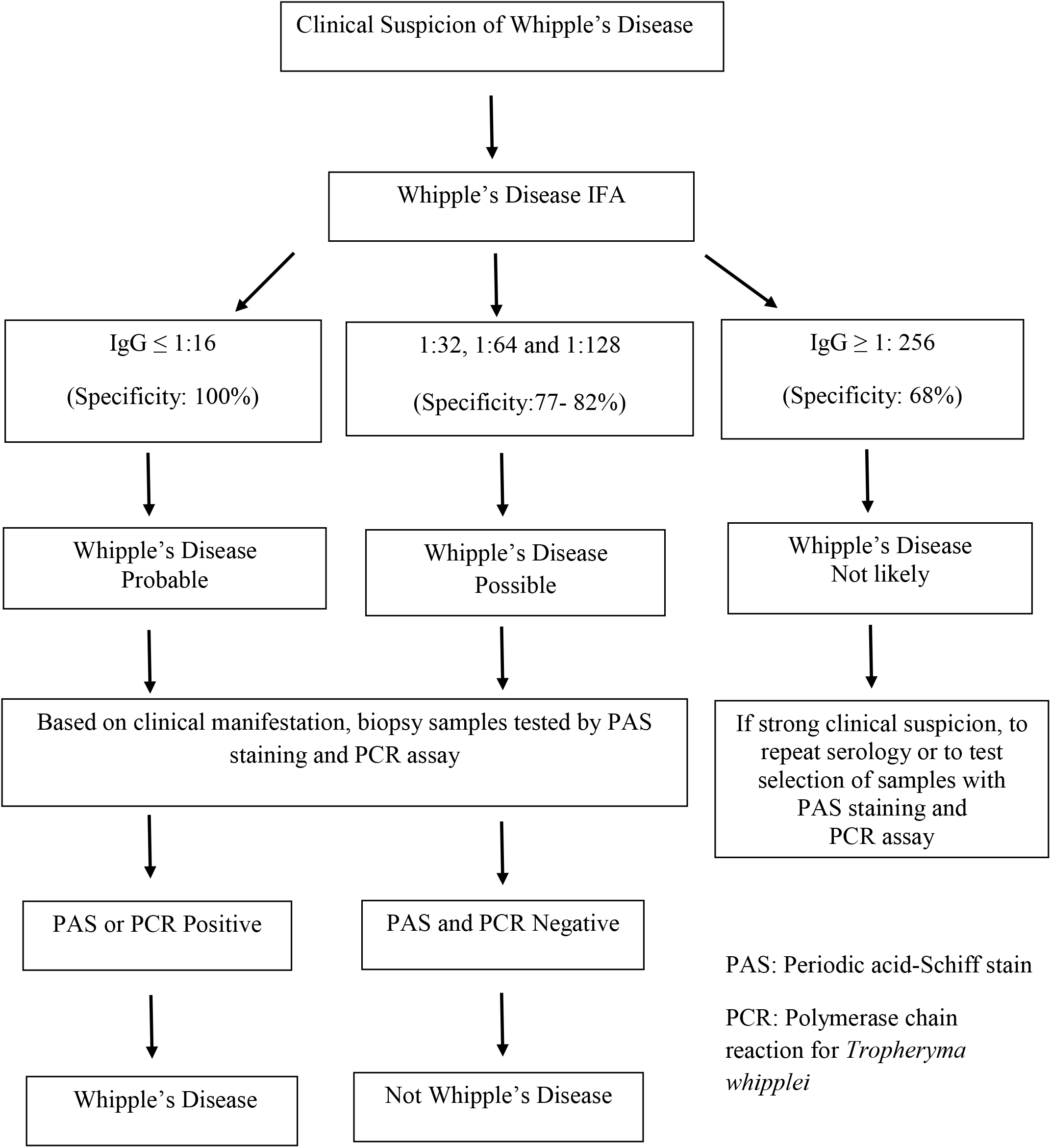
Strategy for Diagnosis and Ruling out Whipple’s Disease Using *Tropheryma whipplei* Indirect Immunofluorescence Assay (IFA)

### IgA IFA

No difference in *T. whipplei* IgA IFA antibody titers was observed between WD cases and controls (*p = 1*.*0*). This antibody isotype does not discriminate between these two groups.

## DISCUSSION

Although small our study appears to confirm the hypothesis that patients with WD under-produce IgG antibody compared with healthy controls. This finding is consistent with previous reports (18,19,22,24). Using ROC analysis, we were able to optimize antibody titer cutoffs and arrive at a potentially useful screening diagnostic assay diagnostically. In patients old enough to have been naturally exposed to *T. whipplei*, those with IgG antibody titers lower than 1:32 are likely to have WD (specificity ∼100%). In a previous French study, when the cutoff titer was set at 1:100, IgG did not discriminate between subjects with and without WD (20). The discrepancy between this result and ours might be explained by the different antibody cutoff titers used as well as possible technical differences in the assays. While these results will need to be replicated and more work needs to be done to determine age- and region-specific rates of WD IgG seropositivity in the general population, we believe that, at least in an Australian context, this assay has the potential to be useful for excluding the diagnosis of WD.

Moreover, while it was less clearly discriminatory than IgG, we also observed differing rates of *T. whipplei* IgM positivity between WD cases and controls. Of note, the pattern of IgM positivity was opposite to that seen in the case of IgG, with IgM detectable at high titer in a significantly *higher* proportion of cases than controls. This finding could suggest a defect in B cell isotype switching from IgM to IgG in WD patients. In our study, only a single WD case (subject #13) appears to have switched from IgM (low titer 1:128) to IgG (high titer 1:4096) (Table1). This subject’s serum sample was collected just days after he underwent open heart surgery, and we speculate that blood transfusions he received might have influenced his antibody readings by detecting donor antibodies as his own.

In contrast to prior studies, we did not find *T. whipplei* IgA to be useful in discriminating between patients with and without WD (21,22). We did not detect serum IgA in any subject from either the case or control group. Secretory IgA in intestinal mucus might correlate to recent exposure to *T. whipplei*, but it would be challenging to measure routinely.

The usual serological response to a primary bacterial or viral infection would involve a conversion of IgM to IgG isotype modulated by T cell cytokines. Seroconversion or a four-fold rise in antibody titer would be expected after about a 10-14-day interval (23). Our serological findings in WD patients were contrary to this. This may be due to a specific genetic predisposition to WD that is associated with human leucocyte antigen (HLA) alleles DRB1*13 and DQB1*06, which may disrupt optimum antigen presentation (25). IL16 gene polymorphisms and other polymorphisms polarize cytokine production towards T-helper-2 (Th2)-cell activity (26-27). This specific genetic predisposition probably explains the inability of switching IgM to IgG antibody isotype. In addition, there appears to be a decrease in Th1 and Th17 activity, poor synthesis of TGF-beta 1 and excess synthesis of IL-10 (19,28-30). Furthermore, bacterial virulence and immunogenicity may be modulated by a bacterial glycoprotein inter-cellular biofilm (18,22,24). This biofilm can prevent the immune system from recognizing the bacterium and impair the switch from IgM to IgG antibody isotype by the patient, a biological mechanism that changes a B cell’s production of antibody from one class to another (18,22,24).

22/22 (100%) of the healthy control group had IgG to *T. whipplei* detected by IFA at a 1:32 serum dilution, or higher, suggesting that the microbe may be widespread in the Australian environment. *T. whipplei* has been detected in 37-66% of waste from sewage plants (4,30-33) and in stool samples of 48% children in Laos (35), 12-26% of sewage plant workers (32) and 1-11% of healthy individuals in France (36). The bacterium is also viable in human feces and saliva (12), supporting the notion of human colonization, probably through fecal-oral transmission. Further work with larger sample populations will be required to fully characterize rates of exposure to *T. whipplei* in relevant communities.

There are several limitations to this study. Only a single strain of *T. whipplei* was used to prepare target antigens for the IFA. Thus, if there are strain-specific differences in antibody affinities, it might have affected our results. Another potential confounder is nonspecific antibody reactivity, especially with antibodies of the IgM isotype. To address this concern, we set the cutoff for IgM reactivity at the relatively high titer of 1:256. Nevertheless, the potential for nonspecific IgM cross-reactivity remains and must be considered in this study’s interpretation. Finally, the absent/low antibody responses we observed in some healthy controls complicate the reliance of absent *T. whipplei* IgG responses as a marker for active WD. We may presume that absence of antibodies in healthy individuals reflects lack of exposure, however the age-specific frequencies of such patterns of serological response remain to be determined.

Despite these limitations, our study demonstrates that *T. whipplei* screening serology is a promising diagnostic modality for this uncommon disease. Given the rarity of WD and the invasive nature of existing diagnostic tests, an inexpensive, non-invasive serological assay with sufficient sensitivity to reliably exclude the diagnosis will be a worthwhile addition to the diagnostic armamentarium.

## Data Availability

Data is stored in Australian Rickettsial Reference Laboratory in hard copy and secure drive. Data related to this study will be stored in agreement with ethics approval.

## Acknowledgments

The authors thank Prof Didier Raoult and Dr Pierre-Edouard Fournier for the French *Tropheryma whipplei* isolate.

They also thank all the patients who were involved in this study and the healthy volunteers from Geelong for assisting with the studies.

Associate Prof Denis Spelman, Associate Prof James Branley, Dr Ashley Watson, Dr Brian Chong, Dr Callum Maggs, Dr Cameron Jeremiah, Dr Carly Hughes, Dr Caroline Bartolo, Dr Christopher Swan, Dr David Andresen, Dr David Foley, Dr David Sheffield, Dr David Sowden, Dr Freya Langham, Dr Hao Yu, Dr Harsha Sheorey, Dr Gabrielle O’ Kane, Dr Hui Yi Ng, Dr Jenny Robson, Dr Katy Lai, Dr Lucy Crawford, Dr Mike Catton, Dr Renjy Nelson, Dr Rob Pickles, Dr Tristan Gibbs, Dr Sandra Jones, Dr Smathi Chong, Dr Su Ann Ho, and Dr Zaal Meher-Homji are thanked for referring their patients to this study.

## Funding

This study was investigators-initiated, supported by the Australian Rickettsial Reference Laboratory.

## Conflicts of interest

All authors: No reported conflicts of interest. All authors have submitted the ICMJE Form for Disclosure of Potential Conflicts of Interest. Conflicts that the editors consider relevant to the content of the manuscript have been disclosed.

## Notes

### Competing Interest Statement

The authors have declared no competing interest.

### Author Declarations

Barwon Health ethics reference number: 19/135

